# Applying Machine Learning to Evaluate for Fibrosis in Chronic Hepatitis C

**DOI:** 10.1101/2020.11.02.20224840

**Authors:** Aravind Akella, Sudheer Akella

## Abstract

Machine learning (ML) is proving to be an appealing analytical tool as modern medicine progresses towards preventative care. Clinical risk prediction models built using ML offer the potential for more effective diagnostic modalities without the need for invasive procedures. With such models, healthcare practitioners may be empowered towards a more preventative approach in management, thus improving clinical outcomes. The management of patients with chronic Hepatitis C is incomplete without considering the presence and extent of liver fibrosis, which is traditionally assessed with biopsy of liver tissue. Although non-invasive testing alternatives to liver biopsy are gaining popularity, they are considered limited due to inadequate accuracy and were designed to be complementary to liver biopsy. In this study, our aim is to build clinical risk models to predict the extent of fibrosis in patients with chronic Hepatitis C using ML algorithms. We developed nine ML algorithms based on an Egyptian cohort dataset, relying only on patient demographics and commonly-obtained serum laboratory values. One of our models was able to evaluate for fibrosis with an accuracy of 0.81, sensitivity of 0.95, and specificity of 0.73. Furthermore, most of our models outperformed many current diagnostic alternatives to liver biopsy for the evaluation of fibrosis in this patient population.

## Introduction

Hepatitis C is a blood-borne infection of the liver caused by the hepatitis C virus (HCV), and is widely regarded as a medical and epidemiological challenge, with detection rates as low as 20% (Spearman et al. 2019). Hepatitis C may present with both an acute and chronic phase in a majority of patients, and include complications such as liver fibrosis and cirrhosis, liver failure, and even liver cancer. Diagnostic evaluation of fibrosis is essential in patients with chronic hepatitis C, as the presence of fibrosis indicates the onset of progressive disease, which may lead to cirrhosis and end-stage liver failure (Perrillo 1997). The extent of fibrosis is best assessed clinically by liver biopsy, which has been traditionally considered gold standard. After histological evaluation, results are classified using the METAVIR scoring system to help monitor the progression of fibrosis. The METAVIR scale ranges from F0, denoting no evidence of fibrosis, up to F4, indicating cirrhosis (see Castera et al. 2015).

Liver biopsy is an invasive procedure with potential clinical complications and results are subject to a variety of statistical errors (Tapper and Lok 2017). More recently, lesser invasive methods of evaluating fibrosis stage have been adopted due to low-cost, ease-of-use, and reproducibility (see e.g., Bedossa et al. 2015; Castera et al. 2015; Patel and Sebastiani 2020). Among them are an array of laboratory-derived indices, such as the ratio of aspartate aminotransferase (AST) to alanine aminotransferase (ALT), the AST-to-platelet ratio index (APRI), and the Fibrosis-4 index (FIB-4) (Bedossa et al. 2015). The performance of these tests to classify clinically significant liver fibrosis can be evaluated by accuracy, sensitivity, specificity, and the area under the receiver-operating curve (AUROC).

Given its scalability and flexibility, the application of machine learning (ML) to disease-specific patient datasets is increasingly being incorporated into predictions for risk stratification, diagnosis and classification, and survival (Ngiam and Khor 2019). Few studies thus far utilized ML algorithms to predict the extent of fibrosis with HCV patient datasets. One study involved data cleansing (Barakat et al. 2019); another established low sensitivities (Hashem et al. 2016); a third study (Wei et al. 2018) combined ML algorithms with existing blood-test-based scoring systems for detection of cirrhosis, but excluded fibrosis prediction. In this study, we apply nine popular ML algorithms to an openly available Egyptian cohort dataset to classify the extent of liver fibrosis. Furthermore, we assess the performance of ML algorithms in addition to clinically available serum laboratory testing.

## Methods

### Dataset

The Egyptian cohort dataset used in this study is downloaded from the repository maintained by the University of California Irvine (UCI) Center for Machine Learning and Intelligent Systems. The dataset contains anonymized diagnosis and treatment information of 1385 adult patients treated in Egyptian hospitals for 18 months. The dataset was presented in a Proceedings paper (Nasr et al. 2017) before being made available on the UCI repository. We used six of the dataset’s 29 features: age, body mass index (BMI), platelet count (Plat), aspartate transaminase in week 1 (AST1), alanine transaminase in week 1 (ALT1), and baseline histological staging (BHS), shown in Table 1. METAVIR fibrosis stage was converted to the corresponding integer, and is denoted as BHS for the purposes of organization.

**Table 1:** Summary of the selected features in the dataset

| Feature | Min. | Max. | Mean | S.D. | 25th<br>Percentile | 50th<br>Percentile | 75th<br>Percentile |
| --- | --- | --- | --- | --- | --- | --- | --- |
| Age | 32 | 61 | 46.3 | 8.8 | 39 | 46 | 54 |
| BMI | 22 | 35 | 28.6 | 4.1 | 25 | 29 | 32 |
| Plat | 93,013 | 226,464 | 158,348 | 38,795 | 124,479 | 157,916 | 190,314 |
| ALT1 | 39 | 128 | 84 | 26 | 62 | 83 | 106 |
| AST1 | 39 | 128 | 83 | 26 | 60 | 83 | 105 |
| BHS | 1 | 4 | - | - | 2 | 3 | 4 |

### Machine Learning Algorithms

Prior to applying ML algorithms for predictive disease analysis, the algorithms require training using a dataset with a known outcome (known as “supervised learning”). A “learned” ML algorithm can then be applied to a dataset with an unknown outcome for disease state prediction. In our study, we trained the ML algorithms with a portion of the Egyptian HCV dataset (which includes liver biopsy results; the gold standard for fibrosis). We then applied the learned ML algorithm to the remainder of the Egyptian HCV dataset to distinguish significant fibrosis (F2, F3, and F4) from early or no fibrosis (F1) and compared the ML algorithm results with those of liver biopsy. The following ML algorithms are used in the present study:

- Logistic Regression
- Naïve Bayes
- Decision Tree
- Random Forest
- Extreme Gradient Boosting
- k-Nearest Neighbor
- Support Vector Machine
- Neural Networks
- Ensemble Method

A detailed description of the ML algorithms was recently presented in an excellent review by Rashidi et al. (2019).

### Applying the Machine Learning Algorithms

Figure 1 shows the steps involved in the application of the ML algorithms to the dataset. No data cleanup was performed as the downloaded dataset appeared clean, without missing or out-of-range values. Three copies of the dataset were cloned, one for each of three experiments (A, B, and C; see step 3 in Figure 1). In experiment A, the observations with non-significant fibrosis (F1) were retained as they are, and those with significant fibrosis (F2, F3, and F4) were grouped together. In experiment B, those with mild fibrosis (F2) were excluded from the group with significant fibrosis, thus combining those with advanced fibrosis (F3) and cirrhosis (F4). In experiment C, the significant fibrosis group included only the group with cirrhosis (F4). Due to class imbalance in experiments A and B, the data was oversampled by randomly duplicating the minority class in these experiments (step 4). Next, variables of the dataset were standardized (step 5), and model parameter (hyperparameter) selection was then performed for each of the ML algorithms using standard libraries (Bergstra 2012) followed by a 10-fold cross-validation wherever appropriate (Pedregosa 2011) (step 6). The dataset was then split into “train” and “test” datasets (step 7), and training the model with the train dataset (step 8) and getting an unbiased evaluation of the final model with the test dataset (step 9).

**Figure 1:**
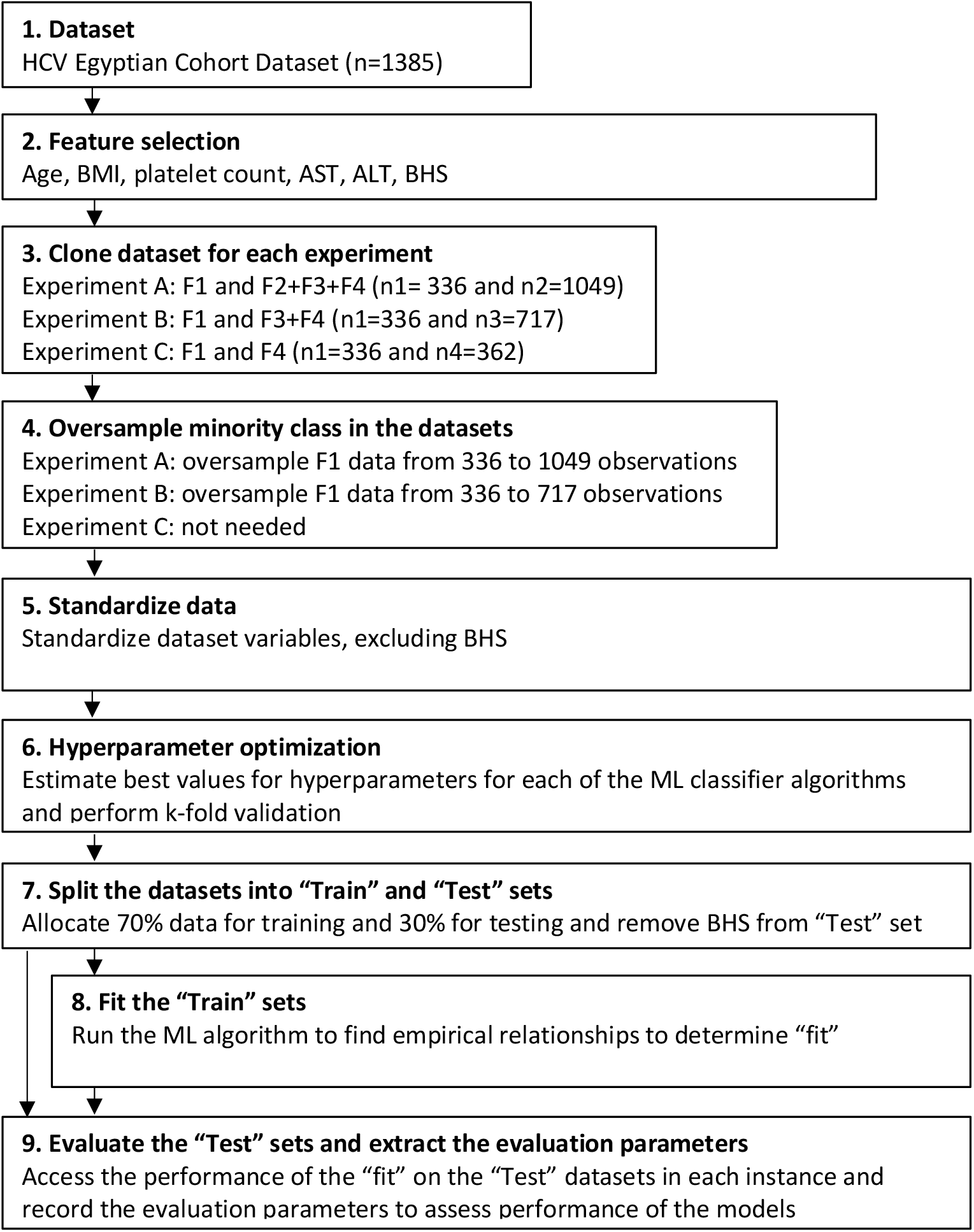
Flowchart of the steps to build and evaluate ML classifier models. “n” indicates the number of observations (rows) in the dataset (see text for details).

### Evaluating the Models

Four parameters (accuracy, sensitivity, specificity, and AUROC) were applied to evaluate the performance of each of our ML models. These parameters can be readily extracted from the confusion matrix computed for each of the models (Sidey-Gibbons and Sidey-Gibbons 2019).

1. Accuracy: the proportion of correct predictions out of the total number of predictions

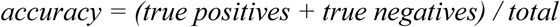
2. Sensitivity: the proportion of correct positive predictions out of real positive instances

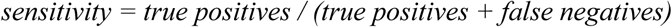
3. Specificity: the proportion of correct negative predictions out of real negative instances

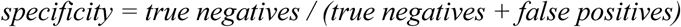
4. AUROC: the estimated probability that a model ranks a randomly chosen positive instance higher than a randomly chosen negative instance.

In addition, the performance of our ML models were compared with the performance of various laboratory diagnostic testing that is currently available.

## Results and Discussion

### Model Evaluation

Figure 2 compares the four evaluation parameters (accuracy, sensitivity, specificity, AUROC) for six of the nine ML algorithms used in our study. Results from three other models are not shown as the values obtained were below 0.5. The evaluation parameters obtained in Experiments A and B are all in the range of 0.60 to 0.96, and generally hovered within 0.34 and 0.64 for Experiment C. Extreme Gradient Boosting (XGB) in Experiment A (XGB-A) performed exceptionally well, with accuracy of 0.81, AUROC of 0.84, sensitivity of 0.95, and specificity of 0.73.

**Figure 2:**
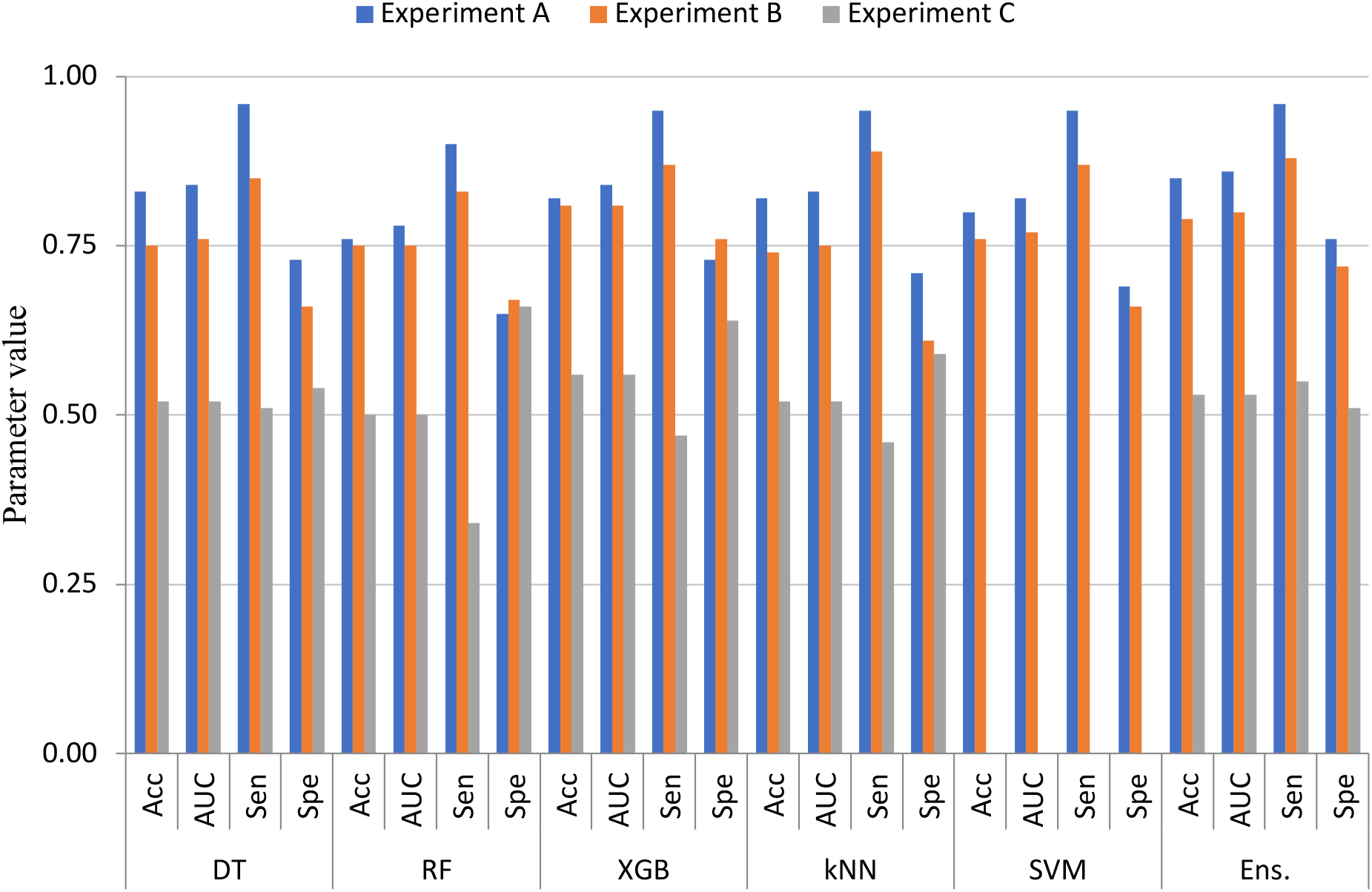
Diagnostic evaluation of the extent of fibrosis in patients in the Egyptian HCV cohort dataset using various machine learning classification algorithms. Results shown are from six machine learning models: DT, Decision Tree; RF, Random Forest, XGB, Extreme Gradient Boosting; kNN, k-Nearest Neighbor; SVM, Support Vector Machine; Ens., Ensemble Method. The evaluation parameters plotted are Acc, accuracy; AUC, AUROC; Sen, sensitivity; Spe, specificity. The results corresponding to Experiment C using SVM were deemed unfit for evaluation and are excluded.

### Comparison with Diagnostic Indices

By comparing the evaluation parameters obtained in our experiments with those corresponding to laboratory diagnostic tests (summarized by Chou and Wasson 2013; Castera et al. 2015), we are able to make direct analysis of the potential of applying ML models over current methods of diagnosis. Table 2 depicts the evaluation parameters for eight laboratory tests along with the evaluation parameters for Extreme Gradient Boosting (XGB) in Experiment A (XGB-A). When compared with laboratory testing methods, our model has both a higher sensitivity (0.95) and AUROC (0.84), as well as a significantly lower negative likelihood ratio (0.07). In addition, the specificity (0.73) and positive likelihood ratio (3.52) of our model fall within the ranges of values corresponding to current diagnostic testing.

**Table 2:** Comparison of evaluation parameters for XGB-A with laboratory diagnostic testing

| Test | Sensitivity | Specificity | AUROC | Positive likelihood ratio | Negative likelihood ratio |
| --- | --- | --- | --- | --- | --- |
| *Summary data from Chou and Wasson (2013)<sup>1</sup>* |  |  |  |  |  |
| Platelet count (<140 to <163) | 0.56 | 0.91 | 0.71 | 6.30 | 0.48 |
| Age-platelet index ($\geq 4$) | 0.70 | 0.70 | 0.74 | 2.30 | 0.43 |
| Age-platelet index ($\geq 6$) | 0.51 | 0.90 | - | 5.10 | 0.54 |
| APRI ($>0.55$) | 0.81 | 0.55 | 0.77 | 1.80 | 0.35 |
| APRI ($\geq 1.5$) | 0.37 | 0.95 | - | 7.40 | 0.66 |
| AST-ALT ratio ($>1$) | 0.35 | 0.77 | 0.59 | 1.50 | 0.84 |
| FIB-4 ($>1.45$) | 0.64 | 0.68 | 0.74 | 2.00 | 0.53 |
| FIB-4 ($>3.25$) | 0.50 | 0.79 | - | 2.40 | 0.63 |
| *Present Machine Learning Study* |  |  |  |  |  |
| Extreme Gradient Boosting (XGB-A) | **0.95** | **0.73** | **0.84** | **3.52** | **0.07** |

<sup>1</sup>Diagnostic performance of laboratory-derived indices for significant fibrosis. Values shown are medians of data gathered from an exhaustive literature search performed by the authors.

## Conclusions and Future Scope

In performing this study, we hoped to demonstrate that clinical risk prediction models built using ML can effectively detect the presence of liver fibrosis in patients with chronic hepatitis C. Our prediction models show improved sensitivity while maintaining similar specificity when compared with various modern laboratory-derived indices that are increasingly used in clinical settings in lieu of liver biopsy. By providing accurate diagnostic evaluation at a lower negative likelihood ratio, our ML-derived models could help clinicians practice preventative medicine by ensuring that less chronic hepatitis C patients with liver fibrosis are undiagnosed and, thus, mismanaged.

That being said, certain considerations should be taken into account in an attempt to further improve the diagnostic promise of our ML-derived models. Firstly, while there are eight confirmed genotypes of HCV that are broken down into further subtypes (Polaris Observatory HCV Collaborations 2017), the Egyptian dataset used in this study is limited to patients diagnosed with chronic hepatitis C due to viral genotype 4a. Furthermore, our models categorically assess for the presence of fibrosis in a binary fashion, whereas pathological analysis of liver tissue is typically classified using the METAVIR scale, which includes five categories to clearly distinguish between various stages of fibrosis. As a result, our models did not assess for cirrhosis; further studies should include data from other HCV cohorts and perhaps consider other disease features to clearly discern the disease state of the HCV patients. Despite the limitations, our study is a significant demonstration of the power of transformative features offered by the ML algorithms in biomedicine to help discern the disease states for clinical use.

Machine learning is increasingly proving to be of transformative value in medical decision-making. The use of ML algorithms as an initial diagnostic tool has the potential to improve the accuracy of disease detection while empowering healthcare practitioners with confidence and preventing potential harm to their patients. The promise of ML may be best realized when algorithms are released as open source, allowing for improved analysis as larger datasets encompassing several cohorts are incorporated, thus providing a self-sustaining model that is adaptable by the medical community.

## Data Availability

The data and code are available on GitHub

https://github.com/aa54/HCV_Egy_ML

## Acknowledgments

We thank Vibhor Kaushik for his assistance in programming using Python.

